# Clinical and Pathological Progression of Awareness Trajectories in Preclinical Alzheimer’s Disease

**DOI:** 10.64898/2026.02.16.26346402

**Authors:** David López-Martos, Gonzalo Sánchez-Benavides, Oriol Grau-Rivera, Rebecca Amariglio, Mark Dubbelman, Jennifer Gatchel, Gad A. Marshall, Ibai Diez, Patrizia Vannini, the A4 Study

**Author notes:** **Corresponding authors:** Ibai Diez, PhD, Center for Inflammation Imaging, 149 13th St, Charlestown, MA 02129, United States, Boston, Massachusetts, 02115, Email; Patrizia Vannini, PhD, Brigham and Women’s Hospital, 60 Fenwood Road, Boston, Massachusetts, 02115, Massachusetts General Hospital, 149 13th Street, Suite 10025, Charlestown, Massachusetts 02129, Email.

## Abstract

Subtle alterations in awareness may emerge in the preclinical stage of Alzheimer’s disease (AD), yet their clinical significance and translational relevance remain unclear. This study aimed to evaluate associations of distinct awareness trajectories with clinical and multimodal AD biomarker measurements in cognitively unimpaired (CU) older adults.

This prospective study analyzed data from the Anti-Amyloid Treatment in Asymptomatic Alzheimer’s (A4) and Longitudinal Evaluation of Amyloid Risk and Neurodegeneration (LEARN) cohorts (∼4.5-year follow-up). Awareness trajectories were defined using a mixed-effects regression model estimating normative longitudinal changes in the Cognitive Function Index (Participant—Study Partner Discrepancy). Based on individual-specific time slopes, participants were classified into three trajectories: stable awareness, heightened awareness (hypernosognosia), and decreased awareness (anosognosia). Study outcomes included the Preclinical Alzheimer’s Cognitive Composite (PACC), Alzheimer’s Disease Cooperative Study (ADCS) Activities of Daily Living Prevention Instrument (ADL-PI), Clinical Dementia Rating (CDR), plasma phosphorylated-tau at threonine 217 (p-tau217), Aβ-PET ([^18^F]-florbetapir), tau-PET ([^18^F]-flortaucipir), and gray matter volume (GMv) via structural magnetic resonance imaging. The associations of awareness trajectories with clinical and multimodal biomarker measurements were evaluated using the general linear model framework, primarily implemented as mixed-effects, including voxel-wise and Braak-stage regional approaches for neuroimaging data. Sequential—longitudinal multimodal neuroimaging mediation analyses evaluated whether regional tau-PET propagation contributed to the emergence of distinct awareness trajectories through downstream GMv loss.

In the full sample (n= 1,643) the mean age was 71.49[±4.72] years, ∼60% female sex, mean education of 16.63[±2.74] years, ∼69% Aβ-PET positive, and ∼27% showing clinical progression on CDR-Global (>0). Compared to stable awareness trajectory (n= 1,325[∼80%]; ∼67% Aβ-PET positive; ∼18% clinical progression), hypernosognosia trajectory (n= 157[∼10%]; ∼68% Aβ-PET positive; ∼36% clinical progression) showed modest clinical implications and limited biomarker associations, including plasma p-tau217, medial temporal tau-PET, and brain structure. In contrast, anosognosia trajectory (n= 161[∼10%]; ∼89% Aβ-PET positive; ∼90% clinical progression) was associated with more adverse outcomes, including steeper cognitive and functional decline, higher risk of progression, greater plasma p-tau217, neocortical tau-PET, and widespread neurodegeneration. Associations between regional tau-PET and awareness trajectories were partially mediated by GMv loss, with sequential Braak-stage II tau-PET effects in hypernosognosia and generalized tau-PET propagation effects extending across Braak-stages II-IV in anosognosia.

These findings suggest that distinct awareness trajectories emerge from stage-specific pathological processes, alongside downstream neurodegenerative mechanisms, reflecting separate clinical consequences. This study identifies anosognosia as a high-risk trajectory across the early stages of the AD continuum, while suggesting that hypernosognosia may reflect both age-related and early AD-related processes.

## Introduction

Anosognosia, the lack of awareness of one’s own neurological condition, manifests in Alzheimer’s disease (AD) as a complete or partial inability to discern cognitive, behavioral, and/or functional deficits.^1–4^ Anosognosia has been well documented in AD, particularly at the stages of dementia and mild cognitive impairment (MCI); showing prevalence estimates ranging from 20% to 80% and greater frequency with disease progression, reflecting variability in sample characteristics, staging, and diagnostic methods.^5,6^ Awareness alterations are typically assessed through clinical judgment, discrepancy between patient and partner reports, or the patient’s subjective report and objective neuropsychological assessment.^6–8^ Several studies have reported associations of anosognosia with increased β-amyloid (Aβ) burden in key AD-related neocortical brain regions;^9,10^ hypometabolism in posterior cingulate cortex (PCC), orbitofrontal cortex (OFC), and hippocampus;^10–12^ reduced frontal, medial parietal, and medial temporal lobe (MTL) brain structures;^13–16^ and altered functional connectivity in key hubs^17^ as well as large-scale brain networks, such as the default mode network (DMN), the salience network (SN), and the frontoparietal network (FPN).^11,12,18^ Clinically, anosognosia has been associated with greater behavioral and neuropsychiatric symptoms,^19–21^ as well as a higher risk of clinical progression across the AD continuum.^9,10^ Despite substantial literature on dementia and MCI stages,^22–30^ research exploring the emergence of altered awareness in the preclinical stage of AD remains limited. Converging evidence suggests that subtle alterations in awareness may emerge alongside subtle cognitive decline, before the onset of clinical impairment.^31,32^

Distinct metacognitive profiles, consistent with heightened awareness (hypernosognosia) and decreased awareness (anosognosia) have been described in cognitively unimpaired (CU) individuals, informing about heterogeneous levels of cognitive awareness across aging and the preclinical stage of AD.^31,33^ Some CU individuals may notice subtle cognitive changes and demonstrate hypernosognosia, underestimating their cognitive performance.^33–36^ This profile resembles subjective cognitive decline (SCD), in which CU individuals express concerns about their cognitive health despite showing no clinical impairment on objective neuropsychological testing.^37,38^ Conversely, other CU individuals may lack insight into subtle cognitive decline, overestimating their cognitive performance.^9,33,36,39–41^ This profile suggests a subclinical form of anosognosia, typically observed in MCI and later stages of AD. Despite their distinct clinical presentations, both hypernosognosia and anosognosia may arise from early AD-related neuropathological changes.^31^ Identifying AD-related changes in awareness could improve early detection and risk stratification; however, further research is needed to determine the clinical, pathological, and neural characterization of subtle alterations in awareness across the preclinical stage of AD.^32^

The objective of this study was to evaluate associations of distinct awareness trajectories with clinical and multimodal AD biomarker measurements in CU older adults. We assessed cognitive and functional performance, clinical progression, plasma phosphorylated tau at threonine 217 (p-tau217), Aβ ([^18^F]-florbetapir) and tau ([^18^F]-flortaucipir) deposition using positron emission tomography (PET), and gray matter volume (GMv) via structural magnetic resonance imaging (MRI). Additionally, sequential and longitudinal multimodal neuroimaging mediation models were employed to investigate whether regional tau PET propagation contributes to the emergence of distinct awareness trajectories through downstream GMv loss in the earliest stages of the AD continuum.

## Materials and methods

### Study population

The present analyses included participants from the Anti-Amyloid Treatment in Asymptomatic Alzheimer’s (A4) and Longitudinal Evaluation of Amyloid Risk and Neurodegeneration (LEARN) cohort studies.^42^ The A4 study is a 4.5-year, placebo-controlled, randomized clinical trial that enrolled CU older adults with evidence of Aβ pathology, as defined by ^18^F-florbetapir PET. The A4 trial evaluated the effect of an anti-Aβ treatment, solanezumab, as compared to placebo, for slowing cognitive decline in preclinical AD [ClinicalTrials.gov NCT02008357].^42^ Solanezumab did not slow cognitive or daily functioning decline in preclinical AD.^43^ The LEARN study consisted of CU participants with non-elevated Aβ levels, serving as a companion observational arm of A4 using same measures and procedures (except treatment). Participants with a Clinical Dementia Rating (CDR) global score of 0, Mini-Mental State Examination (MMSE) score ranging 25-30, and Logical Memory delayed recall score ranging 6-18, were eligible to proceed to Aβ PET imaging with subsequent Aβ status disclosure.^44^ Institutional Review Board (IRB) approval was obtained, and participants provided informed consent in compliance with local IRB and permission to share their de-identified data to advance AD research.^44^

Selection criteria for the present study were based on the availability of both participant and study partner Cognitive Function Index (CFI) measurements at baseline (week 0) and endpoint (week 240). Longitudinal clinical data, including the preclinical Alzheimer’s cognitive composite (PACC), Alzheimer’s disease cooperative study (ADCS) activities of daily living prevention instrument (ADL-PI), and clinical dementia rating (CDR) were selected according to the CFI administration schedule, comprising week 0, 48, 168, and 240. Multimodal biomarker measurements included baseline Aβ PET ([^18^F]-florbetapir), longitudinal plasma p-tau217, longitudinal tau PET ([^18^F]-flortaucipir), and longitudinal GMv. Longitudinal plasma p-tau217 measures were selected to match the clinical schedule, comprising week 0, 48, 168, and 240. Longitudinal tau PET and GMv scans were selected only at baseline (week 0) and endpoint (week 240).

### Clinical measurements

The Cognitive Function Index (CFI) was used to evaluate subjective cognitive concerns.^45^ Participants and their study partners each completed a version of the CFI questionnaire, which asked 15 questions about cognitive concerns relative to 1 year earlier (ranging 0-15, with higher scores indicating greater cognitive concerns). The Participant and Study Partner CFI scores were standardized to the baseline performance of Aβ-negative participants in the LEARN sample to anchor deviations from normative levels (*i.e.*, z-score using mean and standard deviation [SD]), and next subtracted to compute the standardized CFI Discrepancy (CFI Participant – CFI Study Partner), where higher scores indicated increased awareness, reflecting the discrepancy direction of hypernosognosia (subestimation), and lower scores indicated decreased awareness, reflecting the direction of anosognosia (overestimation).

The Preclinical Alzheimer Cognitive Composite (PACC) was used to evaluate global cognitive performance,^46,47^ including the total score on the Free and Cued Selective Reminding Test (FCSRT),^48^ the delayed recall on the Logical Memory test from the Wechsler Memory Scale,^49^ the Digit Symbol Substitution Test from the Wechsler Adult Intelligence Scale–Revised,^50^ and the total score on the MMSE.^51^ The PACC was computed as the arithmetic mean of these four components, standardized to the baseline performance of participants in the LEARN sample (*i.e.*, z-score using mean and SD), with higher scores indicating greater cognitive performance relative to normative levels.

The Alzheimer’s Disease Cooperative Study Activities of Daily Living Prevention Instrument (ADCS ADL-PI) was used to evaluate functional impairment.^52^ ADCS ADL-PI administered to the study partner was used (ranging 0-45, with lower scores indicating greater functional difficulty).

The Clinical Dementia Rating (CDR) scale was used to evaluate global functioning.^53^ In this study, we used the CDR Sum of Boxes score (CDR-SB, ranging 0-18), with higher scores indicating greater impairment in global functioning, and progression on the CDR Global score, which was defined as two consecutive CDR Global scores or a final CDR Global score greater than 0.

### Plasma biomarker measurements

We used already available plasma p-tau217 levels in A4/LEARN studies.^42,54^ Plasma p-tau217 levels were measured using Eli Lilly and Company electrochemiluminescent immunoassay. Sample preparation was automated using Tecan Fluent platform, and detection was performed on the MSD Sector S Imager 600MM.

### Structural MRI and PET data acquisition

Structural MRI, Aβ PET, and tau PET images were available for analysis.^42,55^ For structural MRI imaging, a high resolution 1 mm isotropic 3D T1-weighted sequence was acquired from either General Electrics, Siemens, Phillips Medical Systems or Philips Healthcare scanner. Aβ imaging was acquired using the [^18^F]-florbetapir tracer 50–70 minutes post-injection. The images were reconstructed from four 5-minute frames, except for some sites that utilized a single frame. Tau PET imaging was collected using the [^18^F]-flortaucipir tracer 80 to 110 minutes post-injection. The images were reconstructed from six 5-minute frames, except for some sites that utilized a single frame. Further details regarding data acquisition can be found on the A4/LEARN website (https://www.a4studydata.org/).^44^

### Aβ PET quantification

This study utilized the already computed and available Aβ Centiloid values from the A4/LEARN studies. The original quantification involved the standardization to the Centiloid scale from the mean cortical standardized uptake value ratio (SUVR) using [^18^F]-florbetapir PET imaging, with the whole cerebellum serving as the reference region.

### Structural MRI preprocessing

Structural T1-weighted MRI preprocessing was conducted using FMRIB Software Library (FSL) v6.0.4. The anatomical T1 preprocessing pipeline involved several key steps: initial reorientation to the right-posterior-inferior (RPI) standard space, followed by alignment to the anterior and posterior commissures. Subsequent steps included skull stripping and the estimation of gray matter (GM), white matter (WM), and cerebrospinal fluid (CSF) partial volume using FSL’s FAST tool. Finally, a non-linear transformation was computed to register the individual skull-stripped T1 images to the 2 mm resolution MNI 152 standard template. Additionally, FreeSurfer v6 was used to process structural MRI for PET quantification. To quantify longitudinal changes in GM volume (GMv), the FSL voxel-based morphometry (VBM) approach was employed. The individual subject GM partial volume estimates were registered to MNI 152 standard space using non-linear registration. A left–right symmetric, study-specific GM template was subsequently created by averaging the registered images and flipping them along the x-axis. All native GM images were then non-linearly registered to this study-specific template and modulated to correct for local expansion or contraction induced by the non-linear component of the spatial transformation. Finally, the modulated GM images were smoothed using an isotropic Gaussian kernel with a σ=3.

### Tau PET quantification

The quantification of tau PET data involved a sequential set of established preprocessing steps. First, the PET frames were co-registered and averaged. A subsequent rigid body transformation was applied to align the averaged PET image with the structural MRI image for each participant. The standardized uptake value ratio (SUVR) was then computed using the inferior cerebellum as the designated reference region. To account for tissue atrophy, partial volume correction (PVC) was implemented using the Müller-Gärtner (MG) method, which relies on a FreeSurfer three-compartment model. The corrected images were then transformed into the standard MNI152 space and subjected to spatial smoothing using an isotropic Gaussian kernel with an 8 mm full width at half maximum (FWHM). Finally, a rigorous quality check was performed on the preprocessed data to verify correct processing completion and to assess for any presence of head motion artifacts.

### Regional GMv and tau PET quantification

Mean GMv and tau PET SUVR values were extracted for predefined regions of interest (Braak Stages II, III, and IV) corresponding to the stages of neurofibrillary tau tangles deposition: Stage I (perirhinal); Stage II (entorhinal and hippocampus); Stage III (para-hippocampus, fusiform, lingual, and amygdala); Stage IV (middle/inferior temporal, cingulate, and insula); Stage V (superior/middle/inferior frontal and parietal regions); and Stage VI (pericalcarine, postcentral, cuneus, precentral, and paracentral).

### Definition of awareness trajectories

Awareness trajectories were defined using a mixed-effects regression model that estimated normative longitudinal changes in standardized CFI Discrepancy scores over the study period. The model included age, sex, education, and time, with complete time x covariate interactions. The structure of the models included a random slope and a random intercept, accounting for the effect of time within each participant. For each participant, the individual-specific time slope (*i.e.*, the sum of fixed and random effects) was extracted and standardized to a z-score using the mean and standard deviation of the full sample. To identify meaningful deviations from normative neuropsychological trajectories associated with aging and AD-related decline, a threshold of ±1 z-score on the individual-specific standardized time slope was applied. Participants were thus classified into three awareness trajectories: stable (−1 < z < 1), hypernosognosia (z ≥ 1), and anosognosia (z ≤ −1). The effect of solanezumab treatment on the standardized CFI Discrepancy was assessed using a mixed-effects regression model in the A4 intention-to-treat population, showing no significant treatment effect.

### Statistical analysis

#### Baseline characteristics of the participants

Baseline differences among awareness trajectories, encompassing demographic, genetic, diagnostic biomarkers, and clinical characteristics, were assessed using ANOVAs and chi-squared tests.

#### Clinical outcomes and AD diagnostic biomarkers, including plasma p-tau217 and Aβ PET

Longitudinal clinical and diagnostic biomarker outcomes, including PACC, ADCS ADL-PI partner, CDR-SB, and plasma p-tau217, were evaluated using awareness trajectories as the main predictor of interest (0: Stable; 1: Hypernosognosia; 2: Anosognosia) in mixed-effects regression models adjusted for age, sex, education (except for plasma p-tau217), and time (*i.e.*, days [scaled]), including full time interactions. Similarly, clinical progression, defined as two-consecutive CDR Global scores > 0 or a final CDR Global score > 0, was evaluated using awareness trajectories as the main predictor of interest in a logistic, general linear model (GLM) and Cox proportional hazards (PH) fixed-effects models, both adjusted for age, sex, education, and time (*i.e.*, days [scaled]). Finally, the predictive value of baseline Aβ PET on awareness was evaluated using Centiloids as main predictor of interest and longitudinal measurements of the standardized CFI discrepancy as the outcome in a mixed-effects regression model adjusted for age, sex, education, and time (*i.e.*, days [scaled]), including complete time interactions.

#### Tau PET, as a prognostic AD biomarker, and structural brain morphology

To examine longitudinal changes in regional tau PET and GMv, mixed-effects regression models stratified by early regional Braak stages (II–IV) were used with awareness trajectories as the primary predictor of interest. Models were adjusted for age, sex, education, and time, including full time interactions; with GMv models additionally adjusted for TIV. Considering all the above-mentioned mixed-effects regression models, the structure consistently included a random slope and a random intercept, accounting for the effect of time within each participant, unless, otherwise noted, the structure was decoupled as required for maximum convergence stability. The inclusion of quadratic terms in mixed-effects models was evaluated using R² (Coefficient of Determination) and Akaike Information Criterion (AIC), contrasting complementary information to optimize model performance. Considering ANOVAs, Chi-squared tests, mixed-effects, GLM, and Cox PH, statistical significance was determined as nominal *p* < 0.05. All computational procedures and statistical analyses were performed using R, version 4.2.3, with RStudio, version 2024.04.0.

To identify voxel-level differences in baseline and longitudinal brain imaging metrics among awareness trajectories, a mass univariate voxel-wise analysis was performed using GLMs. Group comparisons were computed using an unpaired t-test within the GLM framework, adjusting for age, sex, and education, with GMv models including further adjustment for TIV. Longitudinal analyses were performed over voxel-wise maps of percentage of change (longitudinal percentage increase in tau PET and percentage decrease in GMv, both relative to baseline values), with additional adjustment for the time interval between baseline and follow-up scans. Results were corrected for multiple comparisons using cluster-wise Monte Carlo simulation with 10,000 iterations to estimate the probability of false-positive clusters, with a statistical significance threshold set at a two-tailed *p* < 0.05. Results were projected onto vertex-wise cortical surface templates and subcortical results were displayed on T1-weighted template slices.

To test the hypothesis that gray matter volume (GMv) loss mediates the association between tau PET and the defined awareness trajectories, a voxel-wise mediation analysis was used. The mediation analysis was executed using GLMs in a three-step process: (i) computing the association between mean regional tau PET SUVR (predictor) and the distinct awareness trajectories (anosognosia or hypernosognosia; outcomes); (ii) identifying voxels where longitudinal GMv change (mediator) is associated with tau PET; and (iii) determining the contribution of tau PET to the prediction of awareness trajectories when the longitudinal GMv change was included in the model. We repeated these analyses for regional Braak stages of interest (II, III, and IV), distinct awareness trajectories, as well as baseline and longitudinal tau PET data. While voxels satisfying all three criteria were classified as fully mediating, the primary focus was on identifying partial mediation; defined by fulfilling criteria (i) and (ii) while observing that the contribution of tau PET became smaller than the contribution of longitudinal GMv change when the mediator was included. This approach allowed us to quantify both the mediated effect through GMv loss and the direct effect of tau PET. All computational procedures and statistical voxel-wise neuroimaging analyses were performed using MATLAB version 2022b.

## Results

A total of 1,643 cognitively unimpaired older adults enrolled in the A4 and LEARN cohorts were included (4.5-year follow-up). Baseline characteristics of the primary study sample are reported in **Table 1** (N = 1,643), whereas baseline characteristics of the subsets with available T1-weighted MRI and tau PET scans (N = 1,125; 332) are provided in **Supplementary Tables 1-2**, respectively.

**Table 1.**
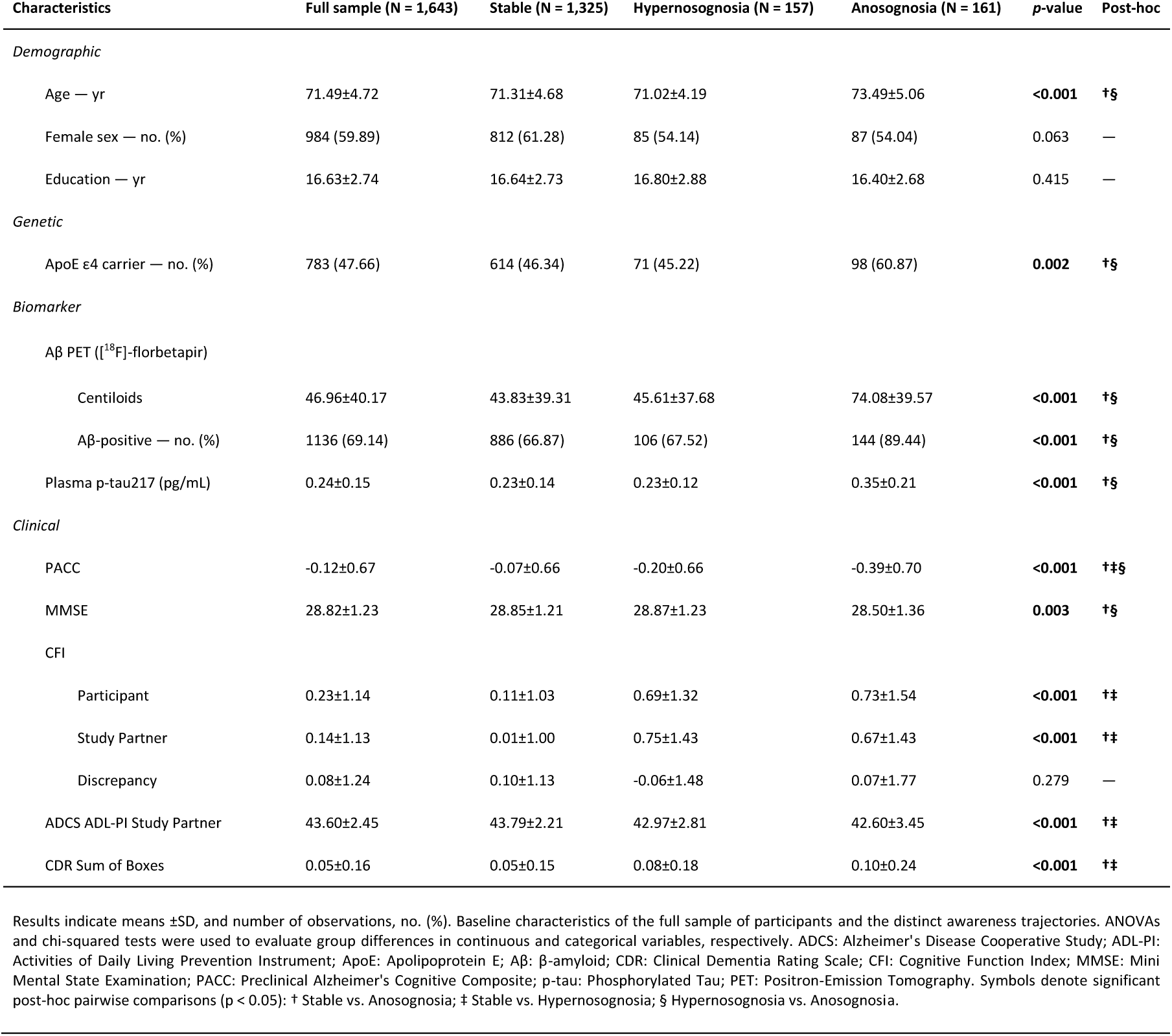
Baseline characteristics of study participants in the primary sample.

### Definition of awareness trajectories

Awareness trajectories were defined using longitudinal mixed-effects modeling of the standardized participant–partner CFI discrepancy, results are presented in **Supplementary Table 3**. Significant differences were observed for the main effect of age (β[95%_CI_]= −0.039 [−0.077,−0.001]), sex (β[95%_CI_]= −0.177 [−0.256,−0.098]), and time (β[95%_CI_]= −0.052 [−0.086,−0.017]), as well as for the time interaction with age (β[95%_CI_]= −0.049 [−0.076,−0.021]). Specifically, older age, male sex, and time from baseline were associated with a lower CFI discrepancy, reflecting lower cognitive awareness, with older age further linked to decline in awareness over time (i.e., overestimation of cognitive performance). Participants were subsequently classified into stable, hypernosognosia, and anosognosia trajectories based on individual-specific standardized time slopes. Most participants were classified as exhibiting a stable awareness trajectory (n = 1,325 [∼80%]; Aβ-PET+ = ∼67%; CDR-Global clinical progression = ∼18%), demonstrating no substantial change in CFI discrepancy over time. This was followed by the anosognosia trajectory (n = 161 [∼10%]; Aβ-PET+ = ∼89%; CDR-Global clinical progression = ∼90%), characterized by a decrease in CFI discrepancy over time, and the hypernosognosia trajectory (n = 157[∼10%]; Aβ-PET+ = ∼68%; CDR-Global clinical progression = ∼36%), characterized by an increase in CFI discrepancy over time. Observed CFI discrepancy z-scores over the 4.5-year period, stratified by awareness trajectory, are shown in **Fig. 1**.

**Fig. 1.**
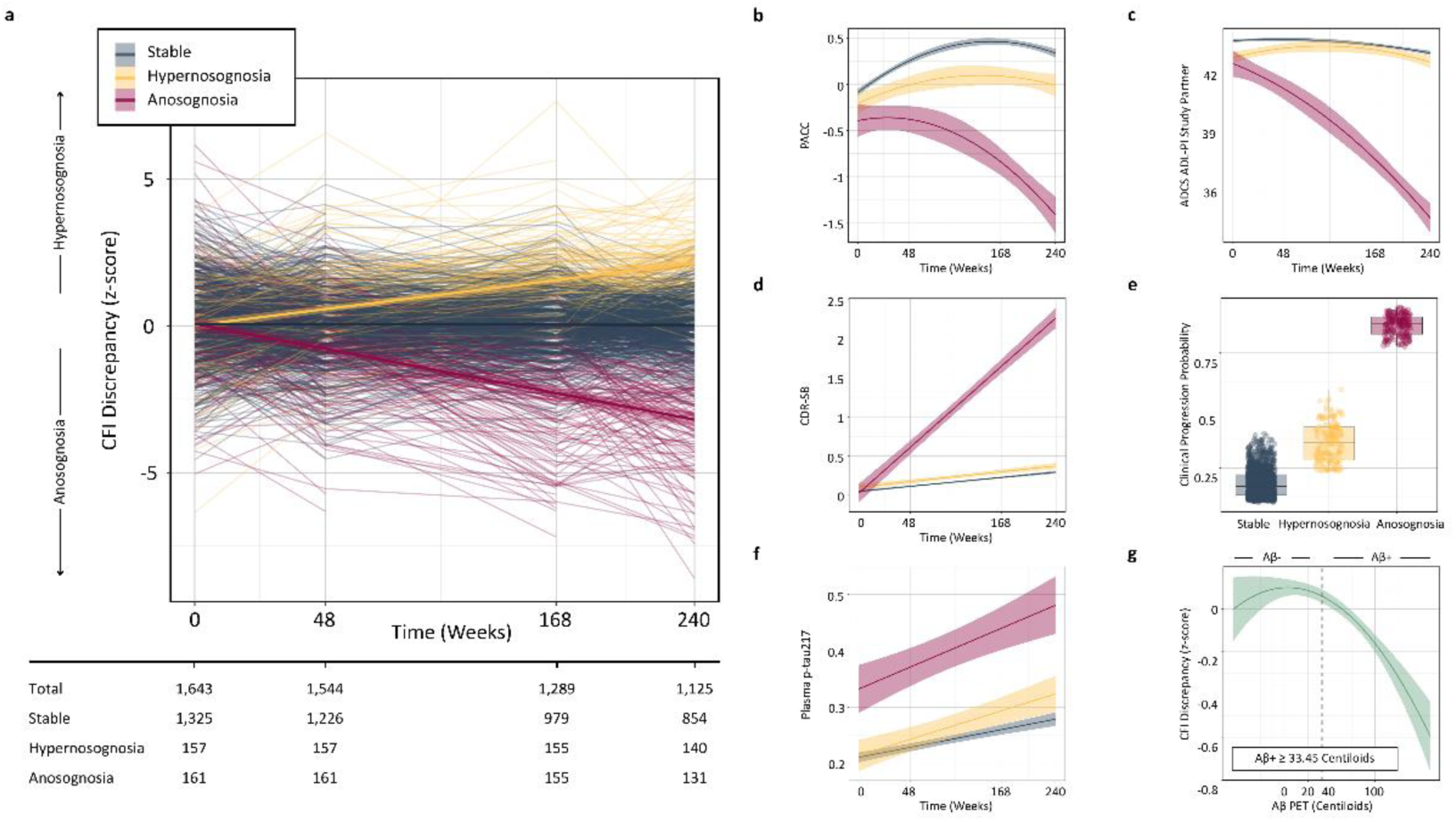
Associations with Clinical and Diagnostic Biomarker Measurements. **a**: Standardized CFI discrepancy (z-scores) over the 4.5-year study period, stratified by awareness trajectories: stable, hypernosognosia, and anosognosia. The accompanying table reports the total number of individuals and the number within each trajectory at every assessment time point (week). **b**: Regression lines showing predicted association of awareness trajectories with PACC, shaded areas represent the 95% CI of regression lines. **c:** Regression lines showing predicted association of awareness trajectories with ADCS ADL-PI Study Partner, shaded areas represent the 95% CI of regression lines. **d**: Regression lines showing predicted association of awareness trajectories with CDR-SB, shaded areas represent the 95% CI of regression lines. **e:** Box plots showing predicted probability of clinical progression associated with awareness trajectories (progression on the CDR Global). **f**: Regression lines showing predicted association of awareness trajectories with plasma p-tau217, shaded areas represent the 95% CI of regression lines. **g**: Regression line showing predicted association of baseline Aβ PET with longitudinal measurements of the standardized CFI discrepancy, shaded areas represent the 95% CI, and the vertical dashed line represents the cut-off for Aβ PET positivity, as defined in the A4/LEARN cohort studies.

### Baseline characteristics of awareness trajectories

Baseline differences across awareness trajectories are summarized in **Table 1**. Significant baseline differences were observed in age (*p* < 0.001), ApoE ε4 (*p* < 0.001), Aβ PET Centiloids (*p* < 0.001), Aβ PET positivity (*p* < 0.001), plasma p-tau217 (*p* < 0.001), PACC (*p* < 0.001), MMSE (*p* < 0.001), CFI reported by both participants (*p* < 0.001) and study partners (*p* < 0.001), ADCS ADL-PI reported by study partners (*p* < 0.001), and CDR Sum of Boxes (CDR-SB; *p* < 0.001). No significant baseline differences were observed for sex, education, or CFI discrepancy. Post-hoc analyses indicated that participants with a hypernosognosia trajectory had lower PACC scores (*p* = 0.030), higher CFI scores reported by both participants (*p* < 0.001) and study partners (*p* < 0.001), lower ADCS-ADL-PI score reported by study partners (*p* < 0.001), and higher CDR-SB score (*p* = 0.013), compared with the stable trajectory. Participants with an anosognosia trajectory had older age (*p* < 0.001), a higher proportion of ApoE ε4 carriers (*p* = 0.001), higher Aβ-PET burden measured by Centiloid units (*p* < 0.001), a greater percentage of Aβ PET positivity (*p* < 0.001), higher plasma p-tau217 concentration (*p* < 0.001), lower PACC score (*p* < 0.001), lower MMSE score (*p* = 0.001), higher CFI scores reported by both participants (*p* < 0.001) and study partners (*p* < 0.001), lower ADCS-ADL-PI scores reported by study partners (*p* < 0.001), and higher CDR-SB score (*p* < 0.001), compared with the stable trajectory. Compared with hypernosognosia, anosognosia was associated with older age (*p* < 0.001), a higher proportion of ApoE ε4 carriers (*p* = 0.007), higher Aβ-PET burden measured by Centiloid units (*p* < 0.001), a greater percentage of Aβ PET positivity (*p* < 0.001), higher plasma p-tau217 concentration (*p* < 0.001), lower PACC score (*p* = 0.008), and lower MMSE score (*p* = 0.007).

### Anosognosia is associated with more adverse clinical and diagnostic AD biomarker outcomes

The associations of awareness trajectories with clinical and diagnostic AD biomarker outcomes are presented in **Fig. 1** and **Supplementary Tables 4-7**. Regarding the PACC, anosognosia was associated with a steeper cognitive decline compared to both the stable trajectory (linear time interaction: β[95%_CI_]= −0.533 [−0.605, −0.461], *p* < 0.001; quadratic time interaction: β[95%_CI_]= −0.082 [−0.132, −0.032], *p* < 0.001) and hypernosognosia (linear time interaction: β[95%_CI_]= −0.439 [−0.533, −0.345], *p* < 0.001; quadratic time interaction: β[95%_CI_]= −0.148 [−0.213, −0.083], *p* < 0.001). Hypernosognosia was associated with lower cognitive performance compared to the stable trajectory (linear time interaction: β[95%_CI_]= −0.094 [−0.165,−0.023], *p* = 0.009; quadratic time interaction: β[95%_CI_]= 0.066 [0.019,0.114], *p* = 0.007), yet no decline relative to its baseline level was observed.

Regarding the ADCS ADL-PI study partner, anosognosia was associated with a steeper decline in daily functioning compared to both the stable trajectory (linear time interaction: β[95%_CI_]= −0.766 [−0.850,−0.682], *p* < 0.001; quadratic time interaction β[95%_CI_]= −0.156 [−0.227,−0.085], *p* < 0.001) and hypernosognosia (linear time interaction: β[95%_CI_]= −0.840 [−0.950, −0.730], *p* < 0.001; quadratic time interaction β[95%_CI_]= −0.104 [−0.196, −0.012], *p* = 0.026). Hypernosognosia showed no association with daily functioning compared to the stable trajectory. Regarding the CDR-SB, anosognosia was associated with a steeper decline in global functioning compared to both the stable trajectory (linear time interaction: β[95%_CI_]= 1.049 [0.962, 1.137], *p* < 0.001), and hypernosognosia (linear time interaction: β[95%_CI_]= 1.047 [0.932, 1.162], *p* < 0.001). Hypernosognosia showed no association with the CDR-SB compared to the stable trajectory. Regarding clinical progression, anosognosia was associated with higher probabilities of clinical progression compared to both the stable trajectory (OR[95%_CI_]= 28.006 [17.616, 46.679], *p* < 0.001; HR[95%_CI_]: 4.173 [3.558, 4.893], *p* < 0.001) and hypernosognosia (OR[95%_CI_]= 11.102 [6.378, 20.056], *p* < 0.001; HR[95%_CI_]: 1.528 [1.219, 1.915], *p* < 0.001). Hypernosognosia was associated with higher probabilities of clinical progression compared to the stable trajectory (OR[95%_CI_]= 2.523 [1.753, 3.604], *p* < 0.001; HR[95%_CI_]: 2.732 [2.153, 3.467], *p* = 0.005). Regarding plasma p-tau217, anosognosia was associated with a steeper longitudinal increase in biomarker concentration compared to both the stable trajectory (linear time interaction: β[95%_CI_]= 0.223 [0.142, 0.304], *p* < 0.001) and hypernosognosia (linear time interaction: β[95%_CI_]= 0.144 [0.041, 0.246], *p* = 0.006). Hypernosognosia was associated with steeper longitudinal increase in biomarker concentration compared to the stable trajectory (linear time interaction: β[95%_CI_]= 0.079 [0.005, 0.154], *p* = 0.035). Regarding Aβ PET, the main effect of Centiloids was non-linearly associated with the standardized CFI discrepancy, showing an inverted U-shaped pattern (linear main effect: β[95%CI]= −0.075 [−0.114, −0.036], *p* < 0.001; quadratic main effect: β[95%CI]= −0.036 [−0.068, −0.004], *p* = 0.030), and the Centiloids x time interaction was linearly associated with decrease in awareness over time (linear time interaction: β[95%CI]= −0.074 [−0.101, −0.047], *p* < 0.001).

### Anosognosia is associated with greater neocortical tau PET propagation and widespread GMv loss

The baseline and longitudinal associations of awareness trajectories with Tau PET and GMv were evaluated using voxel-wise fixed-effects models. Results are presented in **Fig. 2**. Considering baseline analyses, compared to the stable trajectory, anosognosia was associated with higher baseline tau PET deposition across the MTL, inferior temporal, middle temporal, temporoparietal, and frontal regions, along with baseline reduced GMv in MTL, inferior temporal, middle temporal, as well as medial parietal regions. Compared to the stable trajectory, hypernosognosia was associated with higher baseline tau PET deposition in the MTL.

**Figure 2.**
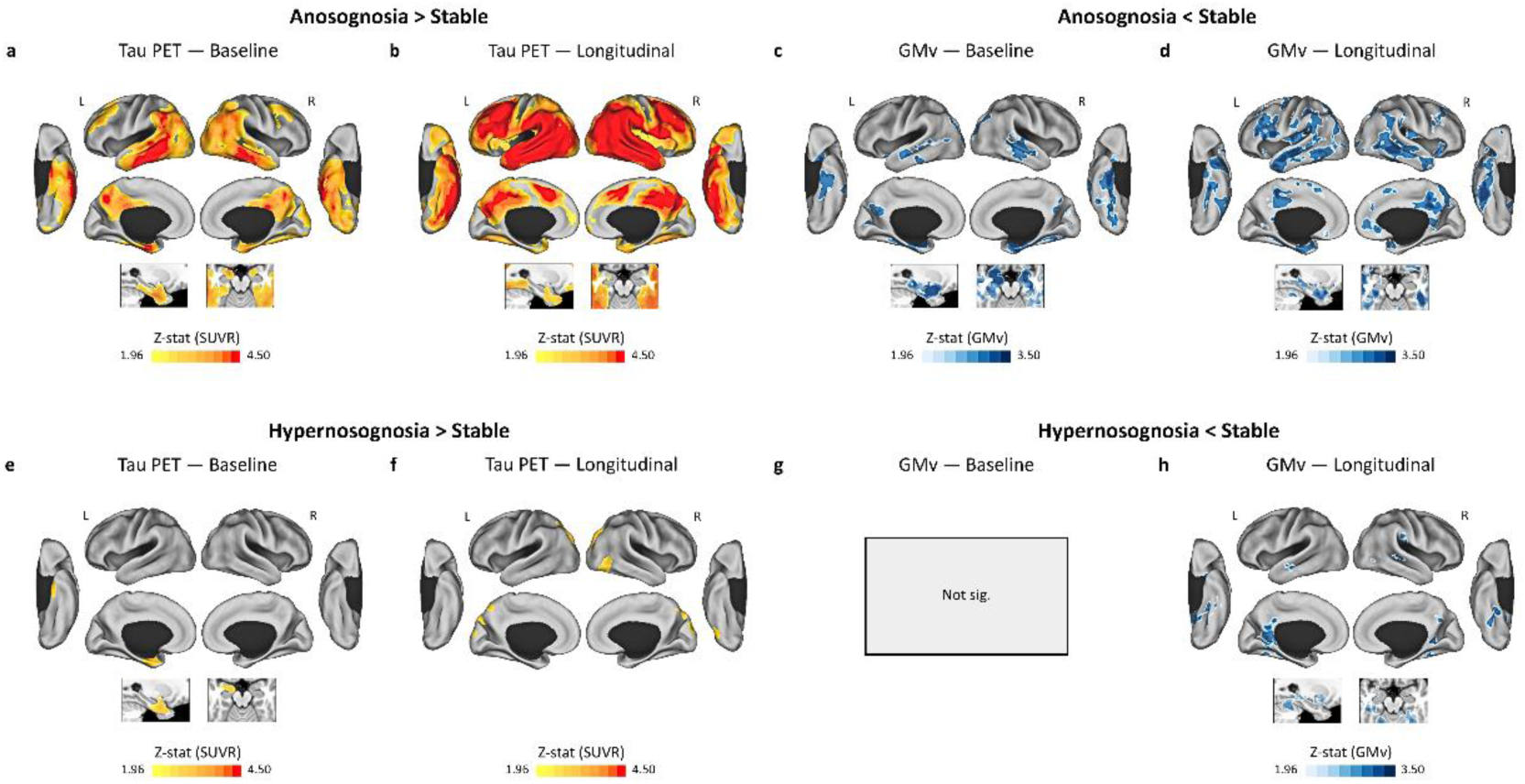
Associations with Prognostic Biomarker and Brain Morphology. Voxel-based tau PET and GMv results are displayed on vertex-wise cortical surface projections and T1-weighted slices. Color bars denote the z-statistics associated with the surface-based projection. Results were corrected for multiple comparisons using cluster-wise Monte Carlo simulation, with 10,000 iterations to estimate the probability of false-positive clusters with a two-tailed *p* < 0.05. **a**: baseline tau PET contrast for anosognosia > stable. **b**: longitudinal tau PET contrast for anosognosia > stable. **c**: baseline GMv contrast for anosognosia < stable. **d**: longitudinal GMv contrast for anosognosia < stable. **e**: baseline tau PET contrast for hypernosognosia > stable. **f**: longitudinal tau PET contrast for hypernosognosia > stable. **g**: baseline GMv contrast for hypernosognosia < stable. **h**: longitudinal GMv contrast for hypernosognosia < stable.

Considering longitudinal analyses, compared to the stable trajectory, anosognosia showed a marked intensification of the baseline pattern. Individuals with anosognosia exhibited greater longitudinal tau PET propagation extending beyond the medial temporal and temporoparietal regions identified at baseline, spreading into a broader, generalized neocortical distribution, including notable change in frontal cortices.

Likewise, longitudinal GMv loss expanded from baseline MTL, inferior temporal, middle temporal, and medial parietal regions toward widespread neocortical regions, with prominent neurodegeneration in frontal but also medial parietal regions. Compared to the stable trajectory, hypernosognosia was associated with longitudinal tau PET propagation across localized brain areas, involving temporo-occipital, and medial parietal regions, along with longitudinal GMv loss across similarly localized regions, mainly within inferior temporal, temporo-parietal, and medial parietal regions.

### Anosognosia and hypernosognosia, voxel-wise and regional comparison

The comparison between anosognosia and hypernosognosia trajectories, considering Tau PET and GMv, was evaluated using voxel-wise fixed-effects and additional mixed-effects models stratified by early regional Braak Stages (II-IV). Results are presented in **Fig. 3** and **Supplementary Table 8**. Compared to hypernosognosia, anosognosia was associated with similar voxel-wise patterns observed when examined against the stable trajectory, though a more restricted spatial extent pattern of tau PET deposition was observed, primarily evidenced by overlapping-contrast baseline differences in MTL, inferior temporal, precuneus, MCC, PCC, mPFC, and dlPFC, and longitudinal differences in inferior temporal, OFC, and insular regions. Similarly, a more restricted pattern of differences in brain morphology was observed, primarily evidenced by GMv overlapping-contrast baseline differences in MTL, inferior and middle temporal, PCC, precuneus, and longitudinal differences in MTL, inferior and middle temporal, ACC, MCC, PCC, precuneus, mPFC, OFC, and insula.

**Figure 3.**
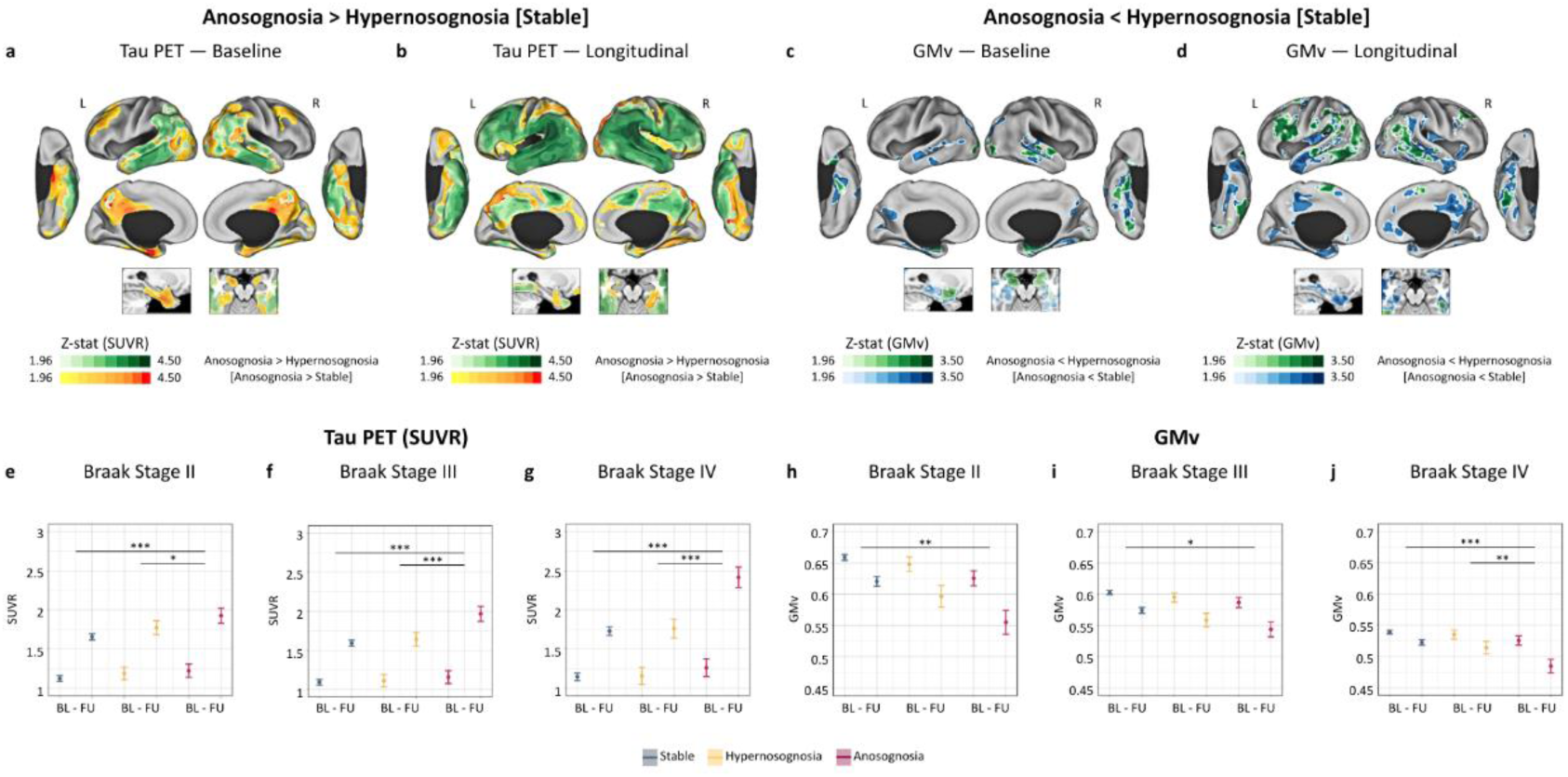
Anosognosia and Hypernosognosia: Voxel-wise and Regional Comparison. Voxel-based tau PET and GMv results are displayed on vertex-wise cortical surface projections and T1-weighted slices. Color bars denote the z-statistics associated with the surface-based projection. Results were corrected for multiple comparisons using cluster-wise Monte Carlo simulation, with 10,000 iterations to estimate the probability of false-positive clusters with a two-tailed *p* < 0.05. **a**: baseline tau PET contrast for anosognosia > hypernosognosia, on top of the baseline contrast for anosognosia > stable. **b**: longitudinal tau PET contrast for anosognosia > hypernosognosia, on top of the longitudinal contrast for anosognosia > stable. **c**: baseline GMv contrast for anosognosia < hypernosognosia, on top of the baseline contrast for anosognosia < stable. **d**: longitudinal GMv contrast for anosognosia < hypernosognosia, on top of the longitudinal contrast for anosognosia < stable. **e-j**: regional analyses across Braak Stages (II-IV) showing estimated marginal means considering time x group interaction corresponding to the predicted associations between awareness trajectories and longitudinal tau PET SUVR (**e-g**) and GMv measurements (**h-j**). Error bars represent 95% confidence intervals of the regression coefficients. Baseline: week 0; follow-up: week 240. Significance codes: *** *p* < 0.001; ** *p* < 0.005; * *p* < 0.05.

Regarding regional analyses of tau PET and GMv by early Braak stages (II-IV). Anosognosia was associated with a steeper longitudinal increase in Braak stage II tau PET burden compared to both the stable trajectory (time interaction: β[95%_CI_]= 0.420 [0.196, 0.644], *p* < 0.001) and hypernosognosia (time interaction: β[95%_CI_]= 0.290 [0.008, 0.572], *p* = 0.044). In contrast, hypernosognosia was not associated with longitudinal change in Braak stage II tau PET burden compared to the stable trajectory. Anosognosia was also associated with a steeper longitudinal GMv loss in Braak stage II compared to the stable trajectory (time interaction: β[95%_CI_]= −0.410 [−0.649, −0.172], *p* = 0.001) but not compared to hypernosognosia. In contrast, hypernosognosia was not associated with longitudinal GMv loss in Braak stage II compared to the stable trajectory. Anosognosia was associated with a steeper longitudinal increase in Braak stage III tau PET burden compared to both the stable trajectory (time interaction: β[95%_CI_]= 0.779 [0.515, 1.043], *p* < 0.001) and hypernosognosia (time interaction: β[95%_CI_]= 0.707 [0.374, 1.039], *p* < 0.001). In contrast, hypernosognosia was not associated with longitudinal change in Braak stage III tau PET burden compared to the stable trajectory. Anosognosia was also associated with a steeper longitudinal GMv loss in Braak stage III compared to the stable trajectory (time interaction: β[95%_CI_]= −0.273 [−0.478, −0.067], *p* = 0.009) but not compared to hypernosognosia. In contrast, hypernosognosia was not associated with longitudinal GMv loss in Braak stage III compared to the stable trajectory. Finally, anosognosia was associated with a steeper longitudinal increase in Braak stage IV tau PET burden compared to both the stable trajectory (time interaction: β[95%_CI_]= 1.085 [0.799, 1.371], *p* < 0.001) and hypernosognosia (time interaction: β[95%_CI_]= 1.045 [0.686, 1.405], *p* < 0.001). In contrast, hypernosognosia was not associated with longitudinal change in Braak stage IV tau PET burden compared to the stable trajectory. Anosognosia was also associated with a steeper longitudinal GMv loss in Braak stage IV compared to both the stable trajectory (time interaction: β[95%_CI_]= −0.546 [−0.768, −0.324], *p* < 0.001) and hypernosognosia (time interaction: β[95%_CI_]= −0.445 [−0.721, −0.169], *p* = 0.002). In contrast, hypernosognosia was not associated with longitudinal GMv loss in Braak stage IV compared to the stable trajectory.

### Neurodegeneration partially mediates the association between regional tau PET and awareness trajectories

Multimodal neuroimaging biomarker analyses evaluated the mediating role of GMv loss on the relationship between regional tau PET (i.e., early Braak stages II-IV) and awareness trajectories. This set of analyses were performed separately by sequential (i.e., expected temporal ordering of key pathological events in AD: baseline tau PET -> longitudinal GMv change -> awareness trajectories) and complete longitudinal models (i.e., longitudinal tau PET change -> longitudinal GMv change -> awareness trajectories). Multimodal mediation was evaluated using GMv voxel-wise fixed-effects models and regional tau PET SUVR measurements. Results are presented in **Fig. 4**. The association between baseline regional tau PET burden and anosognosia across Braak stages II, III, and IV was partially mediated by longitudinal reductions in GMv.

**Figure 4.**
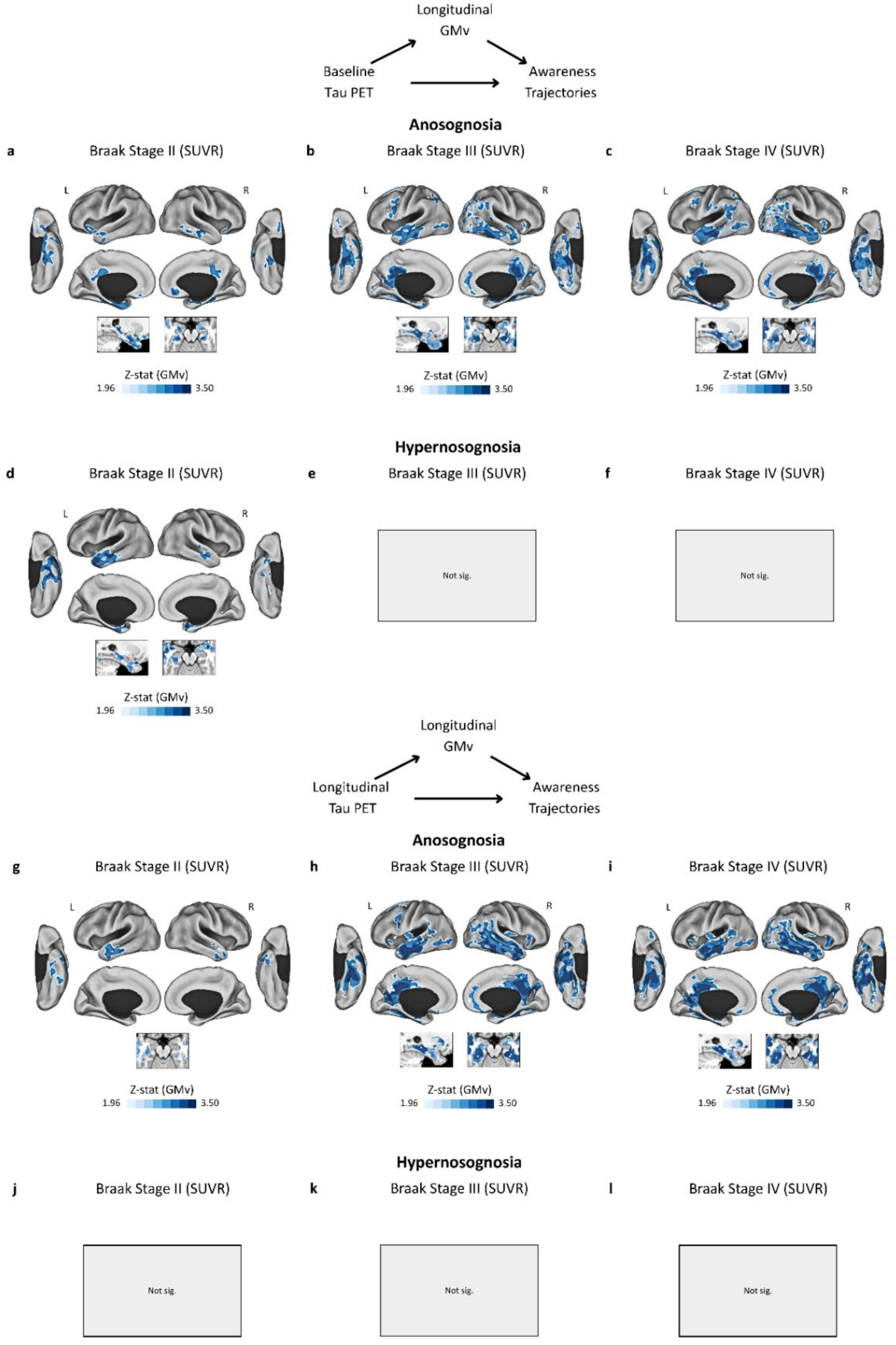
Multimodal Neuroimaging Mediation: Sequential and Longitudinal Analyses. Voxel-based GMv results from the multimodal neuroimaging biomarker mediation models are displayed on vertex-wise cortical surface projections and T1-weighted slices. Color bars denote the z-statistics associated with the surface-based projection. Results were corrected for multiple comparisons using cluster-wise Monte Carlo simulation, with 10,000 iterations to estimate the probability of false-positive clusters with a two-tailed *p* < 0.05. **a-f**: sequential models showing longitudinal GMv contrasts for baseline tau PET SUVR across Braak Stages, indicating associations with anosognosia (**a-c**) and hypernosognosia (**d-f**). **g-l**: longitudinal models showing longitudinal GMv contrasts for longitudinal tau PET SUVR across Braak Stages, indicating associations with anosognosia (**g-i**) and hypernosognosia (**j-l**).

Considering tau PET baseline signal in Braak stage II, the mediating effect of GMv loss was confined to localized regions, including the MTL, inferior temporal, middle temporal cortex, precuneus, PCC, anterior cingulate cortex (ACC), medial prefrontal cortex (mPFC), OFC, and insula. In Braak stages III and IV, the mediating influence of morphological changes extended across the neocortex, encompassing widespread temporal, parietal, and frontal regions. In contrast, the association between baseline regional tau PET and hypernosognosia was only partially mediated by longitudinal reductions in GMv accounting for tau PET SUVR in Braak stage II, where the mediatory effect of GMv loss was restricted to localized regions, including the MTL, as well as inferior and middle temporal regions.

The association between longitudinal regional tau PET propagation and anosognosia across Braak stages II, III, and IV was also partially mediated by longitudinal reductions in GMv. Considering longitudinal tau PET propagation in Braak stage II, the mediating effect of GMv loss was restricted to localized regions, primarily within the inferior and middle temporal cortex. In Braak stages III and IV, the mediating influence of morphological changes expanded substantially across the neocortex, encompassing widespread temporal, parietal, and frontal regions. In contrast, no longitudinal tau PET propagation across Braak stages was associated with hypernosognosia, thereby precluding the mediation analyses for this awareness trajectory.

## Discussion

This study aimed to evaluate the association of distinct awareness trajectories with clinical and multimodal biomarker measurements in CU older adults. Anosognosia trajectory was associated with the most adverse outcomes, including cognitive and functional decline, elevated risk of clinical progression, increased plasma p-tau217, neocortical tau PET burden, and widespread neurodegeneration. Hypernosognosia trajectory showed a less severe profile, with modest clinical implications and limited associations with plasma p-tau217, MTL tau PET, and brain structure. Regional tau-PET associations with awareness trajectories were partially mediated by neurodegenerative mechanisms, with sequential Braak-stage II tau-PET effects for hypernosognosia and generalized tau-PET propagation effects extending across Braak-stages II-IV for anosognosia. These findings showed that distinct awareness trajectories emerge driven by stage-specific pathological processes alongside downstream neurodegenerative mechanisms that give rise to distinct clinical consequences. Together, this study supports cognitive awareness as a clinically meaningful marker for early detection, monitoring, and risk stratification in the preclinical stage of AD.

Awareness trajectories were defined through longitudinal modeling of CFI participant-partner discrepancy, revealing demographic factors shape normative cognitive awareness trajectories, with male sex and older age associated with lower awareness. Individuals exhibiting anosognosia trajectory had higher baseline Aβ PET burden and plasma p-tau217 concentration than those exhibiting hypernosognosia and stable awareness trajectories, whereas those in the hypernosognosia trajectory had similar baseline levels as those in stable trajectory. Anosognosia was associated with cognitive and functional decline, elevated risk of clinical progression, and increasing plasma p-tau217 levels. In contrast, hypernosognosia showed reduced cognitive practice effects, potentially an early indicator of subsequent decline,^56^ no functional impact, modest risk of clinical progression, and modest longitudinal increase in plasma p-tau217. These findings suggest that the anosognosia trajectory represents a high-risk clinical-biological profile, whereas hypernosognosia reflects early neuropathological progression with more limited, yet significant clinical implications.^55,57,58^

Voxel-wise analyses of tau PET and GMv revealed pathological and structural profiles consistent with biomarker staging of AD.^59^ At baseline, anosognosia trajectory exhibited widespread tau PET burden across MTL and neocortex, along reduced GMv in MTL and parietal regions. In contrast, hypernosognosia trajectory showed elevated tau PET burden limited to MTL, with preserved brain structure. Longitudinally, anosognosia trajectory tracked generalized tau PET propagation, primarily across the neocortex, alongside GMv loss extending into frontal and insular regions, whereas hypernosognosia showed modest, localized tau PET spread and GMv loss. Together, these findings showed that anosognosia trajectory reflects AD-related neuropathological changes, whereas hypernosognosia corresponds to less severe pathological changes; which may reflect early AD-related neuropathological changes or primary age-related tau accumulation in the absence of significant Aβ pathology.^59^ These results were primarily consistent with prior reports linking decreased metacognition with reduced cortical thickness in temporal and frontal brain regions in CU older adults.^60^

Direct comparison of hypernosognosia and anosognosia trajectories across voxel-wise analyses of tau PET and GMv showed that anosognosia was associated with greater pathological burden and neurodegeneration. Across neuroimaging modalities, we observed a topographical pattern similar to, but more restricted than, that observed compared to the stable trajectory. This suggested that both awareness trajectories may share a subtle, yet convergent topographical pattern of pathological and structural processes in key brain regions part of the DMN, FPN, and SN. In line, regional analyses of tau PET and GMv revealed descriptive, trajectory-gradient patterns. Anosognosia trajectory exhibited greater tau PET propagation and GMv loss compared to both stable and hypernosognosia trajectories, with hypernosognosia showing longitudinal changes that were intermediate between stable and anosognosia trajectories, but not significantly different from the stable trajectory. In agreement with previous studies, these results showed that while anosognosia trajectory clearly aligns with AD progression, hypernosognosia does not show the same capacity to distinguish AD-related neuropathological changes from those associated with normal aging.^9,11,61^

Multimodal neuroimaging mediation analysis provided key insights into the mechanisms linking regional tau PET, neurodegeneration, and the emergence of awareness trajectories. Sequential modeling showed that GMv loss partially mediated the association between baseline tau PET deposition across Braak stages II–IV and anosognosia. In contrast, mediation was limited to baseline tau PET in Braak stage II for hypernosognosia, reflecting a restricted, yet significant contribution of early-stage tau-related GMv loss to heightened awareness. Complete longitudinal modeling further distinguished awareness trajectories, showing that GMv loss partially mediated the link between longitudinal tau PET propagation across Braak stages II–IV and anosognosia. In contrast, hypernosognosia showed no significant associations with longitudinal tau PET propagation by Braak stages. These results support neurodegeneration as a key downstream pathway through which neurofibrillary tau tangles contribute to clinical manifestations,^62–65^ reflecting pronounced AD-related effects in anosognosia and early effects in hypernosognosia that, however, cannot be definitively attributed to specific AD-related pathological processes.^59^

This study supports non-mutually exclusive scenarios for the early dynamics of cognitive awareness across the AD continuum. First, awareness may follow a non-linear trajectory, emerging as hypernosognosia and gradually progressing to anosognosia.^9,31,33,35,36^ Self-reports of SCD may show an initial increase during aging and preclinical AD, followed by attenuation over time, whereas study-partner reports of SCD continue to rise, driving the participant-partner discrepancy.^31^ Consistently, prior A4 findings showed that PACC correlated more strongly with Study Partner’s CFI than with Participant’s CFI over time, a relationship driven by Aβ burden.^62^ In line, we observed an inverted U-shaped relationship between Aβ PET and the CFI discrepancy consistent with previous cross-sectional studies, indicating a non-linear link between core AD pathology and cognitive awareness.^33,35,36^ Here, we identified an inflection point occurring before the Aβ-positivity threshold, highlighting the emergence of reduced self-awareness early in the AD continuum. Second, hypernosognosia may represent a resilient trajectory in which individuals maintain relatively preserved awareness of cognitive decline over the long term, with discrepancy scores gradually approaching zero as study partners catch up in reporting complaints, without necessarily progressing to anosognosia. In biologically confirmed AD, this pattern could reflect a transient or adaptive metacognitive response, potentially driven by less severe neuropathology or greater resilience mechanisms. Finally, hypernosognosia may also reflect a non-specific trajectory, encompassing individuals with early AD-related changes or primary age-related tauopathy (PART), as well as those with negative biomarkers who report cognitive complaints non-specific for AD, consistent with normal aging, mood symptoms, or functional cognitive disorders.^32,39,41,63–67^

Study limitations should be noted. Definition of awareness trajectories involved inherent trade-offs, while classification was based on the temporal evolution of the CFI, alternative diagnostic approaches and classification schemes could also be applied.^68^ We acknowledge that study partner characteristics may also influence awareness trajectories; however, these factors were not directly incorporated, and this should be considered when interpreting the findings. Similarly, factors such as genetics, lifestyle, or comorbidities were not accounted for in these present analyses. Considering A4 clinical trial results, longitudinal changes in Aβ PET were not assessed.^34^ The A4/LEARN samples consisted primarily of highly educated, non-Hispanic White participants, limiting generalization to diverse populations. Finally, future studies with longer follow-up are needed to determine the long-term significance of awareness trajectories in the preclinical stage of AD.

In conclusion, this study showed that anosognosia trajectory closely tracked clinical and pathological AD progression, highlighting its translational relevance across the early stages of the AD continuum. In contrast, hypernosognosia trajectory showed more limited associations, lacking the same capacity to distinguish normal aging from early AD-related changes. Together, these findings support cognitive awareness trajectories as mechanistically informed markers for early detection, risk stratification, and monitoring of disease progression.

## Acknowledgments

The authors would like to thank all A4 and LEARN participants, their study partners, and the A4 and LEARN Study Teams, including all site principal investigators and staff.

## Data Availability Statement

Data supporting the findings of this study are available through the A4 and LEARN studies. Information on data access can be found at https://www.a4studydata.org/.

