## Supplementary Material for "Clinical and Pathological Progression of Awareness Trajectories in Preclinical Alzheimer’s Disease"

*** Corresponding authors:**

Ibai Diez, PhD

60 Fenwood Road

Boston, Massachusetts, 02115

Massachusetts General Hospital

149 13th Street, Suite 10025

Charlestown, Massachusetts 02129

### Supplementary Material

**Supplementary Table 1.** Characteristics of study participants in the sample with available MRI

| Characteristics | Full sample (N = 1,125) | Stable (N = 854) | Hypernosognosia (N = 140) | Anosognosia (N = 131) | *p*-value | Post-hoc |
|---|---|---|---|---|---|---|
| *Demographic* | | | | | | |
| Age — yr | 71.15±4.42 | 70.88±4.35 | 70.95±4.05 | 73.12±4.79 | **<0.001** | †§ |
| Female sex — no. (%) | 670 (59.56) | 524 (61.36) | 75 (53.57) | 71 (54.2) | 0.091 | — |
| Education — yr | 16.63±2.62 | 16.67±2.57 | 16.71±2.94 | 16.25±2.62 | 0.216 | — |
| *Genetic* | | | | | | |
| ApoE ε4 carrier — no. (%) | 564 (50.13) | 414 (48.48) | 65 (46.43) | 85 (64.89) | **0.001** | †§ |
| *Biomarker* | | | | | | |
| Aβ PET ([$^{18}$F]-florbetapir) | | | | | | |
| Centiloid — units | 48.79±38.78 | 44.91±37.39 | 47.08±37.76 | 75.92±38.26 | **<0.001** | †§ |
| Aβ-positive — no. (%) | 819 (72.8) | 600 (70.26) | 99 (70.71) | 120 (91.6) | **<0.001** | †§ |
| *Clinical* | | | | | | |
| PACC | -0.08±0.68 | -0.01±0.66 | -0.20±0.67 | -0.37±0.72 | **<0.001** | †‡§ |
| MMSE | 28.85±1.22 | 28.89±1.19 | 28.84±1.23 | 28.56±1.38 | **0.014** | † |
| CFI | | | | | | |
| Participant | 2.14±2.03 | 1.85±1.78 | 3.03±2.33 | 3.03±2.59 | **<0.001** | †‡ |
| Study Partner | 1.36±1.90 | 1.06±1.61 | 2.42±2.40 | 2.22±2.33 | **<0.001** | †‡ |
| Discrepancy | 0.77±2.17 | 0.79±1.89 | 0.61±2.65 | 0.82±3.11 | 0.648 | — |
| ADCS ADL-PI Study Partner | 43.64±2.36 | 43.89±2.08 | 42.92±2.83 | 42.75±3.08 | **<0.001** | †‡ |
| CDR Sum of Boxes | 0.05±0.16 | 0.04±0.14 | 0.08±0.19 | 0.11±0.26 | **<0.001** | †‡ |

Results indicate means ±SD, and number of observations, no. (%). Baseline characteristics of the subset of participants with MRI. ANOVAs and chi-squared tests were used to evaluate group differences in continuous and categorical variables, respectively. ADCS: Alzheimer's Disease Cooperative Study; ADL-PI: Activities of Daily Living Prevention Instrument; ApoE: Apolipoprotein E; Aβ: β-amyloid; CDR: Clinical Dementia Rating Scale; CFI: Cognitive Function Index; MMSE: Mini Mental State Examination; PACC: Preclinical Alzheimer's Cognitive Composite; p-tau: Phosphorylated Tau; PET: Positron-Emission Tomography. †: Stable VS Anosognosia; ‡: Stable VS Hypernosognosia; §: Hypernosognosia VS Anosognosia.

**Supplementary Table 2.** Characteristics of study participants in the sample with available tau PET

| Characteristics | Full sample (N = 332) | Stable (N = 240) | Hypernosognosia (N = 48) | Anosognosia (N = 44) | *p*-value | Post-hoc |
|---|---|---|---|---|---|---|
| *Demographic* | | | | | | |
| Age — yr | 71.35±4.57 | 71.30±4.57 | 70.45±3.74 | 72.61±5.15 | 0.073 | § |
| Female sex — no. (%) | 187 (56.33) | 97 (40.42) | 27 (56.25) | 21 (47.73) | 0.110 | — |
| Education — yr | 16.21±2.61 | 16.19±2.60 | 16.62±2.65 | 15.89±2.63 | 0.385 | — |
| *Genetic* | | | | | | |
| ApoE ε4 carrier — no. (%) | 188 (56.63) | 134 (55.83) | 28 (58.33) | 26 (59.09) | 0.893 | — |
| *Biomarker* | | | | | | |
| Aβ PET ([$^{18}$F]-florbetapir) | | | | | | |
| Centiloid — units | 56.07±34.23 | 54.22±33.76 | 53.14±31.67 | 69.34±37.09 | **0.021** | †§ |
| Aβ-positive — no. (%) | 294 (88.55) | 213 (88.75) | 39 (81.25) | 42 (95.45) | 0.100 | — |
| *Clinical* | | | | | | |
| PACC | -0.15±0.71 | -0.05±0.67 | -0.32±0.72 | -0.52±0.71 | **<0.001** | †‡ |
| MMSE | 28.67±1.30 | 28.72±1.28 | 28.55±1.49 | 28.54±1.20 | 0.554 | — |
| CFI | | | | | | |
| Participant | 2.31±2.10 | 2.06±1.90 | 2.91±2.04 | 2.98±2.86 | **0.003** | †‡ |
| Study Partner | 1.50±1.88 | 1.19±1.65 | 2.44±2.10 | 2.13±2.30 | **<0.001** | †‡ |
| Discrepancy | 0.81±2.47 | 0.87±2.20 | 0.47±2.56 | 0.85±3.56 | 0.594 | — |
| ADCS ADL-PI Study Partner | 43.54±2.50 | 43.77±2.33 | 42.81±3.00 | 43.03±2.61 | 0.017 | †‡ |
| CDR Sum of Boxes | 0.05±0.16 | 0.04±0.14 | 0.08±0.19 | 0.11±0.26 | **<0.001** | †‡ |

Results indicate means ±SD, and number of observations, no. (%). Baseline characteristics of the subset of participants with MRI and tau PET. ANOVAs and chi-squared tests were used to evaluate group differences in continuous and categorical variables, respectively. ADCS: Alzheimer's Disease Cooperative Study; ADL-PI: Activities of Daily Living Prevention Instrument; ApoE: Apolipoprotein E; Aβ: β-amyloid; CDR: Clinical Dementia Rating Scale; CFI: Cognitive Function Index; MMSE: Mini Mental State Examination; PACC: Preclinical Alzheimer's Cognitive Composite; p-tau: Phosphorylated Tau; PET: Positron-Emission Tomography. †: Stable VS Anosognosia; ‡: Stable VS Hypernosognosia; §: Hypernosognosia VS Anosognosia.

**Supplementary Table 3.** Definition of Awareness Trajectories

| **Outcome, N [Model]** | **Predictors** | **Std. Coefficient (95% CI)** | ***p*-value** | **$R^2$** | **AIC** |
|---|---|---|---|---|---|
| CFI Discrepancy, 1,643 | | | | 0.016/0.573 | 17845.577 |
| [Mixed-effects: (1 + Time \| Participant)] | | | | | |
| | *Main effects* | | | | |
| | Intercept | 0.069 (0.020, 0.119) | **0.006** | | |
| | Age | -0.039 (-0.077, -0.001) | **0.045** | | |
| | Sex [Female] | -0.177 (-0.256, -0.098) | **<0.001** | | |
| | Education | -0.034 (-0.073, 0.004) | 0.080 | | |
| | Time | -0.052 (-0.086, -0.017) | **0.003** | | |
| | *Time interaction* | | | | |
| | Age | -0.049 (-0.076, -0.021) | **<0.001** | | |
| | Sex [Female] | 0.018 (-0.037, 0.073) | 0.511 | | |
| | Education | 0.015 (-0.013, 0.042) | 0.292 | | |

Mixed-effects regression model evaluating age, sex, education, time, and the corresponding time interactions on the standardized CFI discrepancy. The structure of the model included a random slope and a random intercept, accounting for the effect of time within each participant (1 + time | participant). The number of observations (n) was noted next to the outcome. For categorical variables, the reference level was specified. $R^2$ indicated the marginal/conditional proportion of explained variance and AIC indicated model complexity. AIC: Akaike Information Criteria; CI: confidence interval; $R^2$: Coefficient of Determination; Std: Standardized.

**Supplementary Table 4.** Associations of Awareness Trajectories with Clinical and Diagnostic Biomarker Outcomes

| Outcome, N [Model] | Predictors | Std. Coefficient (95% CI) | *p*-value | $R^2$ | AIC |
|---|---|---|---|---|---|
| PACC, 1,643 | | | | 0.258 / 0.865 | 10162.733 |
| [Mixed-effects: (1 + Time \| Participant)] | | | | | |
| | *Main effects* | | | | |
| | Anosognosia [Stable] | -0.687 (-0.819, -0.555) | **<0.001** | | |
| | Hypernosognosia [Stable] | -0.306 (-0.437, -0.174) | **<0.001** | | |
| | Anosognosia [Hypernosognosia] | -0.381 (-0.557, -0.206) | **<0.001** | | |
| | *Time interaction* | | | | |
| | Anosognosia [Stable] | -0.533 (-0.605, -0.461) | **<0.001** | | |
| | Hypernosognosia [Stable] | -0.094 (-0.165, -0.023) | **0.009** | | |
| | Anosognosia [Hypernosognosia] | -0.439 (-0.533, -0.345) | **<0.001** | | |
| | *Time$^2$ interaction* | | | | |
| | Anosognosia [Stable] | -0.082 (-0.132, -0.032) | **0.001** | | |
| | Hypernosognosia [Stable] | 0.066 (0.019, 0.114) | **0.007** | | |
| | Anosognosia [Hypernosognosia] | -0.148 (-0.213, -0.083) | **<0.001** | | |
| ADCS ADL-PI Study Partner, 1,643 | | | | 0.285 / 0.732 | 26548.656 |
| [Mixed-effects: (1 + Time \| Participant)] | | | | | |
| | *Main effects* | | | | |
| | Anosognosia [Stable] | -0.933 (-1.060, -0.806) | **<0.001** | | |
| | Hypernosognosia [Stable] | -0.066 (-0.191, 0.060) | 0.306 | | |
| | Anosognosia [Hypernosognosia] | -0.867 (-1.035, -0.700) | **<0.001** | | |
| | *Time interaction* | | | | |
| | Anosognosia [Stable] | -0.766 (-0.850, -0.682) | **<0.001** | | |
| | Hypernosognosia [Stable] | 0.074 (-0.009, 0.157) | 0.079 | | |
| | Anosognosia [Hypernosognosia] | -0.840 (-0.950, -0.730) | **<0.001** | | |
| | *Time$^2$ interaction* | | | | |
| | Anosognosia [Stable] | -0.156 (-0.227, -0.085) | **<0.001** | | |
| | Hypernosognosia [Stable] | -0.052 (-0.120, 0.016) | 0.132 | | |
| | Anosognosia [Hypernosognosia] | -0.104 (-0.196, -0.012) | **0.026** | | |
| CDR-SB, 1,643 | | | | 0.433 / 0.669 | 9376.847 |
| [Mixed-effects: (0 + Time \| Participant) + (1 \| Participant)] | | | | | |
| | *Main effects* | | | | |
| | Anosognosia [Stable] | 1.122 (1.030, 1.214) | **<0.001** | | |
| | Hypernosognosia [Stable] | 0.077 (-0.015, 0.168) | 0.100 | | |
| | Anosognosia [Hypernosognosia] | 1.046 (0.924, 1.167) | **<0.001** | | |
| | *Time interaction* | | | | |
| | Anosognosia [Stable] | 1.049 (0.962, 1.137) | **<0.001** | | |
| | Hypernosognosia [Stable] | 0.002 (-0.084, 0.088) | 0.961 | | |
| | Anosognosia [Hypernosognosia] | 1.047 (0.932, 1.162) | **<0.001** | | |
| Plasma p-tau217, 1,422 | | | | 0.405 / 0.932 | -2915.286 |
| [Mixed-effects: (0 + Time \| Participant) + (1 \| Participant)] | | | | | |
| | *Main effects* | | | | |

| | | |
|---|---|---|
| Anosognosia [Stable] | 0.711 (0.569, 0.854) | **<0.001** |
| Hypernosognosia [Stable] | 0.116 (-0.024, 0.255) | 0.104 |
| Anosognosia [Hypernosognosia] | 0.596 (0.408, 0.784) | **<0.001** |
| *Time interaction* | | |
| Anosognosia [Stable] | 0.223 (0.142, 0.304) | **<0.001** |
| Hypernosognosia [Stable] | 0.079 (0.005, 0.154) | **0.035** |
| Anosognosia [Hypernosognosia] | 0.144 (0.041, 0.246) | **0.006** |

Mixed-effects regression model evaluating awareness trajectories on clinical and diagnostic biomarker measurements. Models were adjusted for age, sex, time, and the corresponding time interactions. All models were further adjusted for education, except for plasma p-tau217. The structure of the models included a random slope and a random intercept, accounting for the effect of time within each participant (1 + Time | Participant), unless the structure was decoupled for maximum convergence stability [0 + Time | Participant] + [1 | Participant]). The number of observations (n) was noted next to the outcome. For categorical variables, the reference level was specified. $R^2$ indicated the marginal/conditional proportion of explained variance and AIC indicated model complexity. ADCS: Alzheimer's Disease Cooperative Study; ADL-PI: Activities of Daily Living – Prevention Instrument; AIC: Akaike Information Criterion; CDR-SB: Clinical Dementia Rating – Sum of Boxes; CI: Confidence Interval; PACC: Preclinical Alzheimer Cognitive Composite; p-tau: Phosphorylated Tau; $R^2$: Coefficient of Determination; Std: Standardized

**Supplementary Table 5.** Non-linear Association between Aβ PET and Awareness

| Outcome, N [Model] | Predictors | Std. Coefficient (95% CI) | *p*-value | $R^2$ | AIC |
|---|---|---|---|---|---|
| CFI discrepancy, 1643 | | | | 0.027 / 0.574 | 17815.582 |
| [Mixed-effects: (1 + Time \| Participant)] | | | | | |
| | *Main effects* | | | | |
| | Aβ PET | -0.075 (-0.114, -0.036) | **<0.001** | | |
| | Aβ $PET^2$ | -0.036 (-0.068, -0.004) | **0.030** | | |
| | *Time interaction* | | | | |
| | Aβ PET | -0.074 (-0.101, -0.047) | **<0.001** | | |
| | Aβ $PET^2$ | -0.010 (-0.032, 0.013) | 0.413 | | |

Mixed-effects regression model evaluating baseline Aβ PET on longitudinal standardized CFI discrepancy measurements. The model included linear and quadratic terms for Aβ, was adjusted for age, sex, education, time, and the corresponding time interactions. The structure of the model included a random slope and a random intercept, accounting for the effect of time within each participant (1 + time | participant). The number of observations (n) was noted next to the outcome. $R^2$ indicated the marginal/conditional proportion of explained variance and AIC indicated model complexity. Aβ: β-amyloid; AIC: Akaike Information Criteria; CI: confidence interval; $R^2$: Coefficient of Determination; Std: Standardized.

**Supplementary Table 6.** Associations of Awareness Trajectories with Clinical Progression - General Linear Model

| Outcome, N [Model] | Predictors | Odds Ratios (95% CI) | *p*-value | $R^2$ Tjur | AIC |
|---|---|---|---|---|---|
| Progression on the CDR Global, 1,643 | | | | 0.233 | 1571.017 |
| [GLM: Logistic] | | | | | |
| | *Main effects* | | | | |
| | Anosognosia [Stable] | 28.006 (17.616, 46.679) | **<0.001** | | |
| | Hypernosognosia [Stable] | 2.523 (1.753, 3.604) | **<0.001** | | |
| | Anosognosia [Hypernosognosia] | 11.102 (6.378, 20.056) | **<0.001** | | |

Logistic general linear regression model evaluating the association of awareness trajectories with clinical progression (two consecutive CDR Gobal scores > 0). Models were adjusted for age, sex, and education. The structure of the models was based on fixed-effects only. The number of observations (n) was noted next to the outcome. For categorical variables, the reference level was specified. $R^2$ Tjur indicated the proportion of explained variance and AIC indicated model complexity. AIC: Akaike Information Criterion; CDR: Clinical Dementia Rating; CI: Confidence Interval; $R^2$: Coefficient of Determination; CI: Confidence Interval.

**Supplementary Table 7.** Associations of Awareness Trajectories with Clinical Progression - Cox Proportional Hazards Model

| **Outcome, N [Model]** | **Predictors** | **Hazard Ratios (95% CI)** | ***p*-value** | **C-index (SE)** | **AIC** |
|---|---|---|---|---|---|
| Progression on the CDR Global, 1,643 | | | | 0.705 (0.012) | 10136.69 |
| [Cox PH] | | | | | |
| | *Main effects* | | | | |
| | Anosognosia [Stable] | 4.173 (3.558, 4.893) | **<0.001** | | |
| | Hypernosognosia [Stable] | 1.528 (1.219, 1.915) | 0.005 | | |
| | Anosognosia [Hypernosognosia] | 2.732 (2.153, 3.467) | **<0.001** | | |

Cox Proportional Hazards Model evaluating the association of awareness trajectories with clinical progression (two consecutive CDR Gobal scores > 0). Models were adjusted for age, sex, and education. The structure of the models was based on fixed-effects only. The number of observations (n) was noted next to the outcome. For categorical variables, the reference level was specified. C-index indicated the discrimination capacity and AIC indicated model complexity. AIC: Akaike Information Criterion; CDR: Clinical Dementia Rating; CI: Confidence Interval; C-index: Concordance Index; SE: Standard Error.

**Supplementary Table 8.** Associations of Awareness Trajectories with Regional Tau Propagation and Neurodegeneration by Braak Stages

| **Outcome, N [Model]** | **Predictors** | **Std. Coefficient (95% CI)** | ***p*-value** | **$R^2$** | **AIC** |
|---|---|---|---|---|---|
| Tau PET, 332 | | | | | |
| Braak II | | | | 0.507 / 0.819 | 121.874 |
| [Mixed-effects: (1 + Time \| Participant)] | | | | | |
| | *Main effects* | | | | |
| | Anosognosia [Stable] | 0.243 (0.018, 0.468) | **0.034** | | |
| | Hypernosognosia [Stable] | 0.159 (-0.059, 0.377) | 0.152 | | |
| | Anosognosia [Hypernosognosia] | 0.084 (-0.204, 0.372) | 0.566 | | |
| | *Time interaction* | | | | |
| | Anosognosia [Stable] | 0.420 (0.196, 0.644) | **<0.001** | | |
| | Hypernosognosia [Stable] | 0.130 (-0.079, 0.339) | 0.222 | | |
| | Anosognosia [Hypernosognosia] | 0.290 (0.008, 0.572) | **0.044** | | |
| Braak III | | | | 0.514 / 0.744 | 147.086 |
| [Mixed-effects: (1 + Time \| Participant)] | | | | | |
| | *Main effects* | | | | |
| | Anosognosia [Stable] | 0.166 (-0.059, 0.390) | 0.148 | | |
| | Hypernosognosia [Stable] | 0.051 (-0.166, 0.268) | 0.643 | | |
| | Anosognosia [Hypernosognosia] | 0.114 (-0.172, 0.401) | 0.434 | | |
| | *Time interaction* | | | | |
| | Anosognosia [Stable] | 0.779 (0.515, 1.043) | **<0.001** | | |
| | Hypernosognosia [Stable] | 0.072 (-0.175, 0.319) | 0.567 | | |
| | Anosognosia [Hypernosognosia] | 0.707 (0.374, 1.039) | **<0.001** | | |
| Braak IV | | | | 0.478 / 0.698 | 522.617 |
| [Mixed-effects: (1 + Time \| Participant)] | | | | | |
| | *Main effects* | | | | |
| | Anosognosia [Stable] | 0.221 (-0.011, 0.454) | 0.062 | | |
| | Hypernosognosia [Stable] | 0.022 (-0.203, 0.247) | 0.849 | | |
| | Anosognosia [Hypernosognosia] | 0.200 (-0.098, 0.497) | 0.188 | | |
| | *Time interaction* | | | | |
| | Anosognosia [Stable] | 1.085 (0.799, 1.371) | **<0.001** | | |
| | Hypernosognosia [Stable] | 0.039 (-0.228, 0.306) | 0.772 | | |
| | Anosognosia [Hypernosognosia] | 1.045 (0.686, 1.405) | **<0.001** | | |
| GMv, 1,125 | | | | 0.194 / 0.788 | -3551.667 |
| Braak II | | | | | |
| [Mixed-effects: (1 + Time \| Participant)] | | | | | |
| | *Main effects* | | | | |
| | Anosognosia [Stable] | -0.432 (-0.600, -0.265) | **<0.001** | | |
| | Hypernosognosia [Stable] | -0.139 (-0.300, 0.022) | 0.090 | | |
| | Anosognosia [Hypernosognosia] | -0.293 (-0.509, -0.078) | **0.008** | | |
| | *Time interaction* | | | | |
| | Anosognosia [Stable] | -0.410 (-0.649, -0.172) | **0.001** | | |
| | Hypernosognosia [Stable] | -0.168 (-0.384, 0.048) | 0.128 | | |
| | Anosognosia [Hypernosognosia] | -0.243 (-0.539, 0.054) | 0.108 | | |

| | | | | |
|---|---|---|---|---|
| Braak III | | | 0.210 / 0.847 | -4703.747 |
| [Mixed-effects: (1 + Time \| Participant)] | | | | |
| *Main effects* | | | | |
| Anosognosia [Stable] | -0.303 (-0.469, -0.137) | **<0.001** | | |
| Hypernosognosia [Stable] | -0.146 (-0.306, 0.013) | 0.072 | | |
| Anosognosia [Hypernosognosia] | -0.157 (-0.370, 0.057) | 0.150 | | |
| *Time interaction* | | | | |
| Anosognosia [Stable] | -0.273 (-0.478, -0.067) | **0.009** | | |
| Hypernosognosia [Stable] | -0.146 (-0.332, 0.040) | 0.124 | | |
| Anosognosia [Hypernosognosia] | -0.127 (-0.382, 0.129) | 0.331 | | |
| Braak IV | | | 0.102 / 0.820 | -4950.139 |
| [Mixed-effects: (1 + Time \| Participant)] | | | | |
| *Main effects* | | | | |
| Anosognosia [Stable] | -0.297 (-0.474, -0.121) | **0.001** | | |
| Hypernosognosia [Stable] | -0.090 (-0.259, 0.080) | 0.300 | | |
| Anosognosia [Hypernosognosia] | -0.208 (-0.435, 0.020) | 0.074 | | |
| *Time interaction* | | | | |
| Anosognosia [Stable] | -0.546 (-0.768, -0.324) | **<0.001** | | |
| Hypernosognosia [Stable] | -0.101 (-0.302, 0.100) | 0.324 | | |
| Anosognosia [Hypernosognosia] | -0.445 (-0.721, -0.169) | **0.002** | | |

Mixed-effects regression model evaluating awareness trajectories on regional tau propagation and GMv by Braak Stages (II-IV). Models were adjusted for age, sex, time, and the group x time interaction. Additionally, models predicting GMv were further adjusted for TIV. The structure of the models included a random intercept, accounting for individual baseline differences in the outcome across participants (1 | participant). The number of observations (n) was noted next to the outcome. For categorical variables, the reference level was specified. $R^2$ indicated the marginal/conditional proportion of explained variance and AIC indicated model complexity. AIC: Akaike Information Criterion; CI: Confidence Interval; GMv: Gray Matter Volume; $R^2$: Coefficient of Determination; Std: Standardized; TIV: Total Intracranial Volume.
